# Testing the neural noise account: an investigation of visual temporal precision in Tourette syndrome

**DOI:** 10.1101/2023.03.04.23286794

**Authors:** Hannah R. Slack, Georgina M. Jackson, Stephen R. Jackson

**Author notes:** Correspondence to: Professor Stephen Jackson, School of Psychology, University of Nottingham, NG7 2RD.

## Abstract

Tourette syndrome (TS) is a neurological disorder of childhood onset characterised by the occurrence of vocal and motor tics. The pathophysiology of TS has been linked to dysfunction within cortical-striatal-thalamic-cortical (CSTC) brain circuits and alterations in gamma-aminobutyric acid (GABA) signalling within the striatum. Recently, it has been proposed that increased neural noise, leading to decreased signal-to-noise, may be responsible for dysfunctional information processing in TS. Importantly, increased neural noise could impact all stages of the sensorimotor processing, including sensation, action planning, and action execution, and it currently remains to be determined whether this ‘increased neural noise account’ of TS refers to all aspects of information processing, or is specific to particular stages of information processing. We hypothesised that if TS was associated with decreased signal-to-noise when processing visual stimuli, then this might be reflected in a reduction in the precision of their perceptual timing estimates. To examine this, we investigated the precision of perceptual timing in a group of individuals with tic disorder using a visual temporal-order-judgement (TOJ) task.

## Introduction

Tourette syndrome (TS) is a neurological disorder of childhood onset characterised by the occurrence of vocal and motor tics (Leckman, 2002). Tics are repetitive, stereotyped behaviours that occur with a limited duration (Cohen et al., 2013). Motor tics can be simple or complex, ranging from simple repetitive movements to coordinated action sequences. Verbal tics can consist of repetitive sounds, words or utterances, the production of inappropriate or obscene utterances, or the repetition of another’s words. Tics occur in bouts, typically many times in a single day, and are the most common form of movement disorder in children (Cohen et al., 2013).

The volitional nature of tics is debated (Cavanna et al., 2017). While tics are often referred to as involuntary movements (e.g., Ganos et al., 2015), others have argued that tics are voluntary, and occur in response to so-called ‘premonitory urges’ (Leckman et al., 2006). Thus, the majority of individuals with TS report that their tics are often preceded by premonitory urge phenomena (PU) that are described as uncomfortable feelings or bodily sensations that occur prior to the execution of a tic, and are experienced as a strong urge for motor discharge (Cohen et al., 2013). Individuals who experience PU report that these experiences are more bothersome than their tics, that expressing their tics give them relief from, and temporarily abolishes, their PU, and that they would not exhibit tics if they did not experience PU. For this reason, it has been proposed that PU should be considered as the driving force behind the occurrence of tics, and that tics are a learnt response to the experience of PU (Cavanna et al., 2017; Leckman et al., 2006). Within this view, tics are seen as voluntary responses to aversive sensory stimulation.

Individuals with Tourette syndrome often report that many of their tics can be voluntarily suppressed, but that it can be uncomfortable and stressful to suppress tics, and that the urge to tic increases during suppression. Therefore, an alternative perspective on the relationship between PU and tics is that PU occur primarily in circumstances where tics are suppressed, or their execution is deferred (Jackson et al., 2011). Specifically, a distinguishing feature of movements associated with urges, as distinct from involuntary actions, is that urges are chiefly associated with actions that cannot be realized immediately and must be held in check until an appropriate time when they might be released (Jackson et al., 2011).

The voluntary/involuntary nature of tics is important insofar as it has also been argued that volitional actions are accompanied by a distinctive subjective experience, and as a result, feel different from physically similar involuntary movements (Ganos et al., 2015). It is proposed that in Tourette syndrome, the presence of tics may result in the blurring of the boundary between voluntary and involuntary movements, resulting in ‘impaired perception of the different subjective experiences accompanying these two distinct kinds of action’ (Ganos et al., 2015). Importantly, it has been proposed that Tourette syndrome may be particularly associated with increased neural noise leading to decreased signal-to-noise during information processing (Münchau et al., 2021).

Consistent with this view, Adelhöfer and colleagues recently demonstrated, using EEG recording techniques, that so-called 1/f noise was increased in individuals performing a sensorimotor integration task (Adelhöfer et al., 2021). However, the increased 1/f noise observed in this study was not related to clinical measures of tic severity, and as the effects of increased neural noise could impact different stages of the sensorimotor task, i.e., sensation, action planning, or action execution, it remains to be determined whether increased ‘neural noise’ in Tourette syndrome refers to all aspects of information processing or is more specific, e.g., to sensation, or, motor planning, or motor execution.

In support of the proposal that increased neural noise might contribute to deficits in action execution, there is evidence that individuals with TS are less precise on a motor timing tasks than those without TS (Martino et al., 2019). Similarly, in support of the proposal that increased neural noise might contribute to deficits in action planning in TS, there is evidence that individuals with TS may have less precise (i.e., noisier) forward sensory model estimates, resulting in less accurate movements. Finally, in support of the proposal that increased neural noise might contribute to alterations in sensory processing in TS, there is indirect evidence to indicate that individuals with TS experience heightened sensitivity to external stimuli in all five senses and that this may arise due to a breakdown in sensory gating mechanisms (Buse et al., 2016; Patel et al., 2014). It is noteworthy, however, that sensory thresholds are typically within the normal range in TS, indicating that alterations in patients’ perceived sensation most likely arise due to altered central processing of sensory stimuli (Patel et al., 2014), and that individuals with TS have been shown to exhibit paradoxically reduced interoceptive awareness in interoceptive detection tasks, such as heartbeat tracking (Ganos et al., 2015; Rae et al., 2019).

To further examine this issue, we investigated the precision of perceptual timing in a group of individuals with tic disorder using a visual temporal-order-judgement (TOJ) task that required individuals to make an *unspeeded* judgement on each trial with respect to which of two visual stimuli first appeared on the computer display. We hypothesised that, if TS was associated with decreased signal-to-noise when processing visual stimuli, then this would be reflected in a reduction in the precision of their perceptual timing estimates.

## Method

### Design

The current study used a mixed factorial design. The between-subject independent variable was the participants’ tic disorder diagnosis status. The within-subject independent variable was the stimulus onset asynchrony (SOA) between the visual stimuli presented on each trial in a temporal order judgement task. Specifically, SOA referred to the temporal asynchrony between the onset of two visual stimuli (white squares measuring 150 × 150 pixels. presented to the left or right of a central fixation point) presented on each trial. The dependent variable was participants’ temporal precision in judging which of the two squares was presented first on each trial. This was calculated in the following manner. First, for each observer the proportion of ‘right-first’ responses was calculated for each SOA and the best-fitting straight line was then computed. Second, the slope of this line was calculated. Finally, the *just noticeable difference - JND* (i.e., the smallest interval that is needed to reliably indicate temporal order) was then computed. This is calculated by subtracting the SOA required to achieve 75% performance from that required to achieve 25% performance and then dividing this value by 2.

### Participants

Participants were recruited for online participation in this study through the UK charity Tourettes Action, through our existing database of research participants with a tic disorder, and via social media posts. As outlined below, participants’ demographic information was first collected online using *Qualtrics* and then participants were re-directed to *Pavlovia* to complete the behavioural task. Participants who did not correctly provide their demographic information or go on to complete the behavioural task were omitted from the data analysis. A total of 128 individuals correctly provided their demographic details and went on to complete the behavioural task. All participants provided informed consent to take part in the study and the study was approved by the ethical review committee of the School of Psychology, University of Nottingham (**S1455**). The median age was 18 ± 3.5 (inter-quartile range) years (95% CI: 17 – 51 years). 45 participants reported having a diagnosed tic disorder or reported having motor or vocal tics currently. 83 participants reported having no tic disorder diagnosis and did not exhibit tics. The median age for the TS (tic disorder) group was 28 ± 21 years, whereas the median age for the CS (control - no tic disorder) group was 18 ± 1 years.

### The Temporal-Order-Judgement Task

A visual TOJ task was used to measure participants’ ability to accurately perceive the temporal asynchrony between two visual stimuli. The task was implemented using PsychoPy software and ran online using Pavlovia (Peirce et al., 2019). Visual stimuli consisted of two white squares, each measuring 150 × 150 pixels. Participants first provided their demographic information via an online survey hosted on Qualtrics before being redirected to the temporal order judgement task. Upon opening the temporal order judgement task, participants saw a black instructions screen with details on how to complete the task. Onscreen text also instructed participants to turn off their mobile phone and/or TV to prevent any distractions. Participants could begin the trials at any time by pressing the spacebar. Participants first completed 5 practice trials to familiarise themselves with the task, before progressing to the main experiment trials.

On each trial, participants first saw a white fixation cross displayed at the centre of the screen, which remained visible for a variable interval (*range* = 0.3-1.0 seconds). After this, two white squares were presented on the left and right sides of the screen. The onset time of each square relative to the other differed on each trial, such that a square could either precede, succeed, or arrive at the same time as the other square. Across the task, the SOA between the two squares ranged between from -140ms to 140ms increasing in steps of 20ms. A negative SOA indicated that the left square onset prior to the right square. Whereas a positive SOA demonstrated that the opposite was true. When the SOA was set to 0ms, both squares appeared at the same time.

Participants responded by pressing the “S” key on their keyboard if they believed that the left square had appeared prior to the right square. Alternately, participants pressed the “K” key if they believed that the right square was shown before the left square. Responses could only be made once both squares were visible. Participants had 2s in which to make their response. If no response was made during the trial, participants were reminded of the task instructions via text which was presented onscreen for 1s. Each trial lasted approximately 2s. An inter-trial interval was presented for 100ms, during which a black screen was displayed. The task consisted of 150 trials that ran in a pseudo-random order with 10 trials per SOA level. The full procedure lasted approximately 5-minutes.

## Results

### Slope values

Simple regression analyses were conducted to determine the slope of the best-fitting straight line fit for each observer. Figure 1 presents the best-fitting straight line fits of TOJ data for the CS and TS groups.

**Figure 1:**
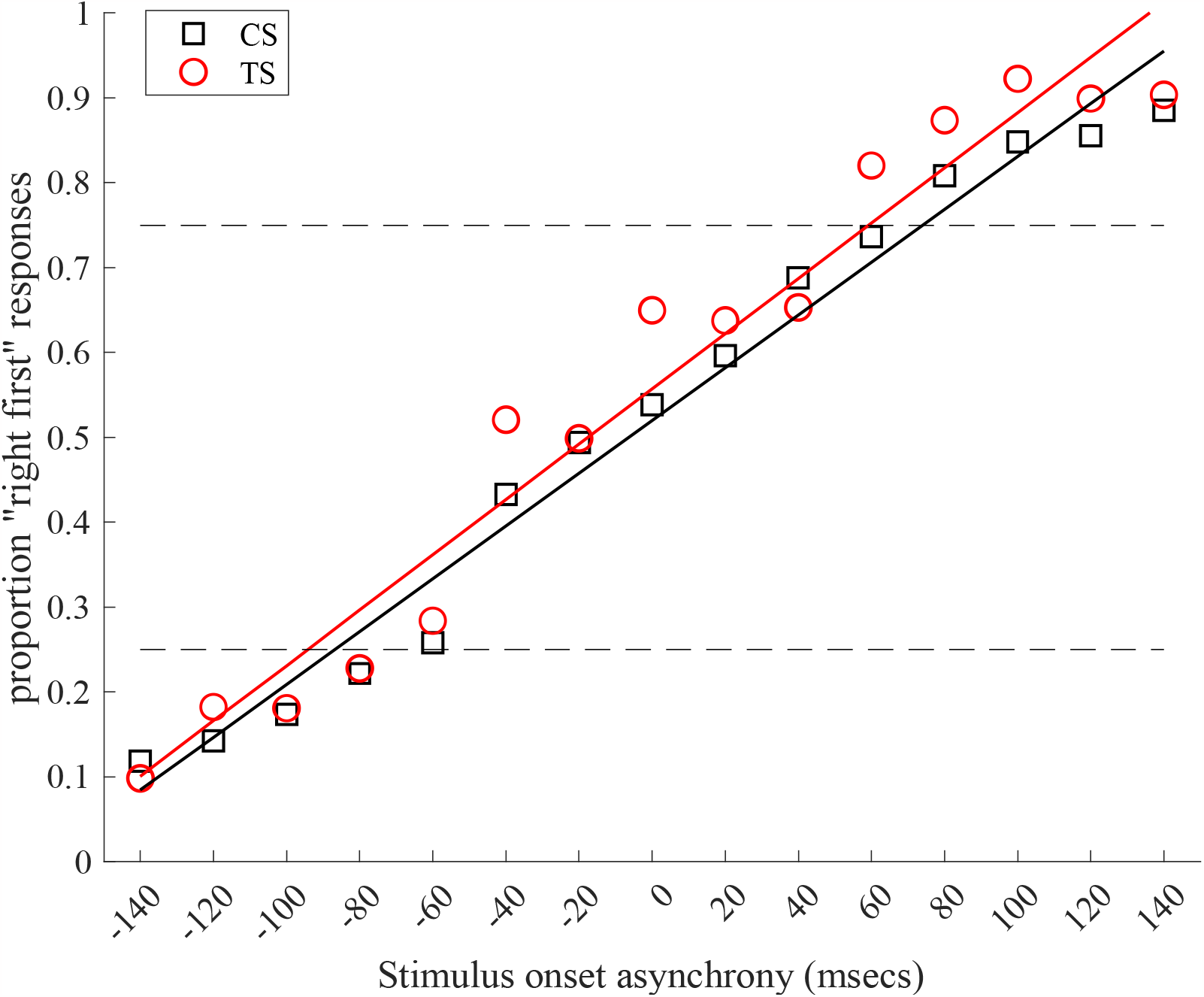
Illustrates the mean proportion of ‘right-first’ responses as a function of SOA, and the best-fitting straight line for each group. The horizontal broken black lines indicate the 0.25 and 0.75 values used in the calculation of the JND.

The analyses revealed that the mean slope value for the CS group was 0.52 ± 0.1 and for the TS group was 0.55 ± 0.16. Between group comparison of mean slope values was conducted by computing the effect size for the standardised difference between the means for each group (Hedges’ G), and by testing for a statistical difference between the means using an independent group t-test. These analyses confirmed that the standardised difference between the means was small (Hedges G = -0.23, 95% CI: -0.69 to 0.23) and that the difference was not statistically significant (*t-*value = -1.0, *p*-value = 0.32).

**Table 1:** Presents results of statistical analyses of between-group differences in means for the slope of the best-fitting straight line fit and just-noticeable-difference (JND) measurements.

| Variable | CS group | | TS group | | Effect size | Low 95% CI | High 95% CI | $t$ | $P$ |
| --- | --- | --- | --- | --- | --- | --- | --- | --- | --- |
|  | Mean | SD | Mean | SD |  |  |  |  |  |
| Slope | 0.52 | 0.1 | 0.55 | 0.16 | -0.23 | -0.69 | 0.23 | -1 | 0.32 |
| JND (ms) | 71 | 10.6 | 68.2 | 13.4 | 0.24 | -0.12 | 0.61 | 1.32 | 0.19 |

### JND values

The mean JND value for the CS group was 71.0 ± 10.6ms and for the TS group was 68.2 ± 13.4ms. Between group comparison of mean JND values was conducted by computing the effect size for the standardised difference between the means for each group (Hedges’ G), and by testing for a statistical difference between the means using an independent group t-test. These analyses confirmed that the standardised difference between the means was small (Hedges G = 0.24, 95% CI: -0.12 to 0.61) and that the difference was not statistically significant (*t*-value = 1.32, *p*-value = 0.19).

### Effects of age

As recruitment for the study was completed online, the ages of the CS and TS groups were not particularly well matched. In fact, as was noted above, the median age for the TS (tic disorder) group was 28 ± 21 years whereas the median age for the CS group was 18 ± 1 years. To ensure that differences in age were not contributing to the null finding reported above for JND values, we conducted a stepwise multiple regression analysis in which we first entered age then group into the analysis. This regression analysis confirmed that neither age (coefficient = -0.09, *t*-value = -0.88, *p*-value = 0.38) nor group (coefficient = 2.83, *t*-value = -1.3, *p*-value = 0.2) were significant predictors of JND value. Thus, while the ages of the observers were not well matched, there is no indication that age was a factor in producing the null effect.

## Discussion

This study was motivated by the proposal that, as a consequence of increased neural noise, TS is associated with decreased signal-to-noise during information processing (Münchau et al., 2021). We hypothesised that, if TS was associated with decreased signal-to-noise when processing visual stimuli, then this would be reflected in a reduction in the precision of their perceptual timing estimates. To investigate this we conducted a simple visual temporal-order-judgement (TOJ) task comparing the temporal precision of the TOJ judgements of a group of individuals with TS against a control group of individuals without tics. The results of this study clearly indicated that there was no evidence to support the hypothesis that temporal precision was reduced in TS. Specifically, we found that there was no difference in the mean JNDs of the TS and control groups or in the mean slopes of the best-fitting straight line fits. These results are discussed below.

The proposal that TS is associated with increased neural noise leading to altered signal-to-noise during information processing follows from a consideration of sensorimotor tasks that often involve complex perception-action binding (Münchau et al., 2021). Increased neural noise could influence all stages of sensorimotor processing, including sensory processing, action planning, and action execution. Alternatively, increased neural noise might primarily effect only one of these stages.

The proposal that sensory processing mechanisms might be dysfunctional in TS arises from two main sources of evidence. The first is that individuals with TS will very often report uncomfortable bodily sensations (PU) that precede their tics and are experienced as a strong urge for motor discharge (Cohen et al., 2013). For this reason, PU have been considered the driving force behind the occurrence of tics, and their presence has often been interpreted as evidence for sensory dysfunction in TS (Cavanna et al., 2017; Cox et al., 2018; Leckman et al., 2006). Second, Individuals with TS report experiencing sensory hyper-sensitivity which is characterised as an increased or prolonged response to sensory stimuli that would not normally be experienced as aversive by a neurotypical observer (Belluscio et al., 2011; Isaacs & Riordan, 2020; Isaacs et al., 2022). Thus, a recent study reported that 86.7% of individuals with chronic tic disorder exhibited higher scores on a self-rated measure of sensory hyper-sensitivity (Sensory Gating Inventory - SGI) than neurotypical controls (Isaacs et al., 2022). It should be noted that, whereas PU phenomena have often, but not always, correlated with tic severity scores (Cavanna et al., 2017), and are transient phenomena that are closely time-locked to the onset of tics, it is argued that sensory hyper-sensitivity is a constant increased awareness of external and internal sensory stimulation, that is uncorrelated with measures of tic severity or PU (Isaacs & Riordan, 2020). One explanation for sensory hyper-sensitivity is that it represents an imbalance between mechanisms responsible for *sensitisation* (i.e., increased response to stimuli following repeated exposure) and mechanisms responsible for *habituation* [i.e., reduced response to stimuli following repeated exposure] (Isaacs et al., 2022). Consistent with this proposal, it has been shown that individuals with TS exhibit impaired sensory gating insofar as they demonstrate abnormal habituation to sensory stimulation (Buse et al., 2016; Patel et al., 2014).

We argue that neither the occurrence of PU phenomena or the presence of sensory hyper-sensitivity provide compelling evidence for an impairment in sensory processing in tic disorders like TS. First, PU do not precede *all* tics, even in individuals who report experiencing very strong PU. Specifically, many individuals exhibit tics, the execution of which they are unaware (e.g., eyeblink or facial tics), and these tics rarely, if ever, are preceded by PU. Instead, it has been argued that PU phenomena, characterised by a strong urge-for-action, occur when actions are consciously suppressed or their execution delayed (Jackson et al., 2011). Within this view, PU arise not through any abnormality in sensory processing but are instead a normal response to the withholding or suppressing an action, and can be observed with respect to a wide variety of actions, e.g., suppressing a yawn, suppressing blinking, withholding urination, etc. (Brown et al., 2017; Jackson et al., 2011). Second, despite the repeated demonstration of sensory hyper-sensitivity in TS, several studies have demonstrated that sensory detection thresholds are typically within the normal range in TS (Patel et al., 2014), indicating that alterations in sensitivity to perceptual stimuli in TS most likely arise as a result of altered high-level, interpretative, perceptual processing rather than through dysfunctional sensory processing. Consistent with this view, the findings of the current study indicate that there is no evidence to indicate that the temporal precision of visual TOJ judgements made by individuals with TS is any different from those made by neurotypical individuals who do not have a tic disorder. Furthermore, this null effect cannot be explained as resulting from group differences in age.

Does this null finding cast doubt on the ‘Neural noise account of TS’? We would suggest not. Tic disorders such as TS are associated with the occurrence of tics, i.e., unwanted movements and vocalisations, and while tics might well have external or internal environmental triggers, this need not imply that sensory processing is dysfunctional. It remains entirely plausible that increased neural noise, leading to reduced signal-to-noise, is primarily associated with the neural circuits directly linked to motor preparation and movement execution that are implicated in the occurrence of tics.

TS has been particularly associated with dysfunction within ‘motor’ cortical-striatal-thalamic-cortical (CSTC) brain circuit that is implicated in the selection of movements and in habit learning (Albin & Mink, 2006), and with significant reduction of gamma-aminobutyric acid (GABA) inhibitory inter-neurons within the striatum (Kalanithi et al., 2005). It is likely that such a reduction in inhibitory signalling within the striatum would lead to an increase in motor ‘noise’ and the generation of unwanted movements. Consistent with this proposal, studies that have investigated the effects of striatal disinhibition in rodents and non-human primates, have consistently demonstrated that the localised administration of a micro-injection of GABA_A_-antagonists reliably produces tic-like movements (Bronfeld, Israelashvili, et al., 2013; Bronfeld, Yael, et al., 2013). That said, a detailed consideration of the evidence for increased motor ‘noise’ in TS is beyond the scope of this paper which has focused on investigated the temporal precision of visual perception in TS.

## Data Availability

All data produced in the present study are available upon reasonable request to the authors

## Acknowledgments

Hannah R. Slack is supported by an Economic and Social Research Council (ESRC) PhD studentship [Grant number: ES/P000711/]. Stephen R. Jackson is supported by grants from the Medical Research Council (MRC) [T032588], Tourettes Association of America, Tourettes Action (UK), and the NIHR Nottingham Biomedical Research Centre.

## Notes

### Competing Interest Statement

The authors have declared no competing interest.

### Author Declarations

The School of Psychology Ethics Committee of the University of Nottingham gave ethical approval for this work (S1455).

